# Mucin VNTR haplogroup associations with idiopathic pulmonary fibrosis are not independent of rs35705950

**DOI:** 10.64898/2026.11.11.26356288

**Authors:** Carl J Reynolds, Richard J Allen, Katherine A Fawcett, Edward J Hollox, Louise V Wain, Anna Peljto, David A Schwartz

## Abstract

**Background:** The *MUC5B* promoter variant (rs35705950) is the strongest common genetic risk factor for idiopathic pulmonary fibrosis (IPF). *MUC5B* and *MUC5AC* contain variable number tandem repeat (VNTR) regions. Long-read sequencing resolves *MUC5AC* into three SNP-tagged haplogroups (H1, H2, H3) differing in repeat length, and *MUC5B* into two (H1, H2) differing in lineage but not length. The tags correlate with rs35705950, so whether a haplogroup effect is distinguishable from the promoter variant is unclear.

**Methods:** We assigned *MUC5AC* and *MUC5B* haplogroups from imputed chromosome 11 array data in 771 European-ancestry IPF Job Exposures Study participants (385 cases, 386 controls). Logistic regression adjusted for age, smoking, principal components and centre, with and without rs35705950. The same models were fitted in TOPMed (1,961 cases, 3,816 controls).

**Results:** *MUC5AC* H3 and *MUC5B* H1 were each associated with IPF (H3 OR 1.63, 95% CI 1.26–2.09, p=1.8×10⁻⁴; *MUC5B* H1 OR 0.71, 0.52–0.98, p=0.038). Both attenuated on adjustment for rs35705950 (H3 OR 1.10, 0.83–1.45, p=0.52; *MUC5B* H1 OR 0.82, 0.58–1.15, p=0.25). A *MUC5AC* H2 association persisted after adjustment (OR 0.69, 0.51–0.94, p=0.017). In TOPMed the H3 findings were consistent with those from IPFJES (unadjusted OR 1.62, 1.47–1.78, p<10⁻¹⁵; adjusted 0.94, 0.84–1.05, p=0.26), but the rs35705950-adjusted H2 association did not replicate: the estimate was null overall (OR 0.99, 0.88–1.11, p=0.87) and in men and women separately (1.00, 0.87–1.16; 0.97, 0.79–1.19). The *MUC5B* H1 and H2 haplogroups could not be tagged in TOPMed.

**Conclusions:** The *MUC5AC* H3 and *MUC5B* H1 associations are not independent of rs35705950. The *MUC5AC* H2 association seen here did not replicate. VNTR haplogroup does not appear to contribute to IPF risk independently of rs35705950.

**Take-home message:** In a dedicated IPF case-control study, the *MUC5AC* H3 and *MUC5B* H1 associations were not separable from the *MUC5B* promoter variant rs35705950. A *MUC5AC* H2 association that did survive adjustment here was not replicated in a larger independent sample.

## Introduction

Idiopathic pulmonary fibrosis (IPF) is a progressive and frequently fatal fibrotic lung disease (Raghu et al., 2022). The strongest common genetic risk factor is a gain-of-function promoter variant in the mucin gene *MUC5B* (rs35705950; minor allele frequency ∼9% in Europeans), whose minor (T) allele confers an odds ratio of approximately 4 per allele (Seibold et al., 2011; Allen et al., 2017) and explains an estimated 2–11% of variation in IPF risk against an overall heritability of 28% (Moll et al., 2023). It increases *MUC5B* expression in the bronchiolar epithelium and is thought to predispose to fibrosis through mucociliary dysfunction (Seibold et al., 2011; Hancock et al., 2018) or epithelial cell stress (Dobrinskikh et al., 2023; Bridges et al., 2025).

*MUC5B* and *MUC5AC* lie within the 11p15.5 mucin cluster and encode gel-forming mucins with large central variable number tandem repeat (VNTR) regions. Plender et al. used long-read sequencing to define three *MUC5AC* haplogroups (H1 short, H2 medium, H3 long) and two *MUC5B* haplogroups (H1 less common, H2 common), with tagging SNPs (tSNPs) allowing assignment in short-read data (Plender et al., 2024). Both are phylogenetic clades from putatively neutral non-coding sequence, but the *MUC5AC* clades also differ in repeat length (H3 is ∼670 amino acids longer than H1), whereas the MUC5B protein clades do not.

There are biological grounds to ask whether haplogroup matters beyond rs35705950. The VNTR encodes the serine- and threonine-rich backbone carrying the O-glycans that set mucus viscoelasticity, and MUC5AC and MUC5B are the principal determinants of mucociliary clearance (Hill et al., 2022; Abrami et al., 2024). VNTR structure could therefore act on that mechanism through protein architecture rather than expression. Length is not the only route: clades differ in other architectural features and may carry regulatory or other coding variation. At *MUC5B*, where length is essentially invariant in Europeans (Plender et al. report expanded VNTRs in seven haplotypes, all of African ancestry), any effect must be of that kind, which is the reason to test the *MUC5B* haplogroups despite their shared repeat length. This has not been tested directly. In the *All of Us* phenome-wide scan no *MUC5AC* haplogroup tag survived multiple-testing correction, and the *MUC5B* locus was represented by rs35705950 rather than a haplogroup tag (Plender et al., 2024). A genome-wide tandem-repeat study in IPF reported an associated repeat at 11p15.5 that lost significance once rs35705950 was included, but its catalogue was restricted to repeat units of ≤20 bp, so it indexed a shorter intergenic repeat rather than the *MUC5AC* (24 bp) or *MUC5B* (87 bp) coding VNTRs, and it did not test the haplogroup classification (Oketch et al., 2025). We therefore ask, in IPFJES (the IPF Job Exposures Study), a dedicated IPF case-control study (Reynolds et al., 2023), how much haplogroup association, if any, is independent of rs35705950.

## Methods

### Study population

IPFJES is a multicentre UK case-control study of men, recruiting from 21 hospitals in England, Scotland, and Wales (Reynolds et al., 2023). IPF cases were diagnosed at multidisciplinary team meetings by 2011 ATS/ERS/JRS/ALAT guidelines; recruitment ran from February 2017 to October 2019. Controls were participants without IPF from the same centres, frequency-matched for age. All participants provided written informed consent. Ethical approval was granted by the East Midlands – Nottingham 1 Research Ethics Committee (REC 17/EM/0021; IRAS 203355). Of 902 IPFJES participants with clinical data, 829 also had typed-array and imputed chromosome 11 genotypes with complete covariates. One centre (n=31) enrolled cases only, so its centre indicator predicted case status perfectly and the corresponding coefficient could not be estimated (complete separation); that centre was excluded, leaving 798. Because genetic ancestry is the principal confounder here and adjustment alone may not remove it, analyses were then restricted to participants of self-reported white ethnicity, who form a single genetic cluster: 25 of the 27 excluded lie beyond 3 SD of the white-ethnicity mean on PC1. This gave an analytical sample of 771 participants (385 cases, 386 controls) used throughout, with principal components recomputed within it.

### Genotyping and imputation

Participants were genotyped on the Biobank Affymetrix Axiom array (GRCh37). As previously described (Chin et al., 2026), sample-level quality control removed individuals with low genotyping call rate (<95%), high heterozygosity, sex mismatches, duplicates, and high relatedness. Imputation used the TOPMed reference panel (GRCh38); imputed variants with r²<0.5 or minor allele count ≤3 were excluded.

### Haplogroup assignment

We used a two-step procedure. First, 17 *MUC5AC* tSNPs were extracted from our GRCh38 imputed data using PLINK v2: 16 of the 92 tags listed in Plender et al. Supplementary Table S4, plus rs35779873 from their Table S6. Second, one concordance SNP was chosen per haplogroup as the primary tag, the H3 and H2 tags entered as allele-dosage counts and H1 serving as the reference: H3 by rs28542750 (r²=1.0) and H2 by rs879201833 (r²=0.888, the best available H2 tag), with H1 as reference (the most common haplogroup, r²=0.899 for rs769768817). The remaining 14 tSNPs were retained as a cross-check on those three (Supplementary Table S1). Tagging r² values are from Plender et al. Supplementary Table S4, except rs35779873, whose r² is from their Table S6. Allele orientation was resolved separately for each tSNP and recorded with its evidence: from the risk allele given in Plender et al. Table S6 where listed, and otherwise by matching the counted-allele frequency in our controls to the published haplogroup frequency. Plender et al. name the two concordance tSNPs in their main text but do not give their alleles, so those were oriented by frequency; rs35705950 was oriented by its established risk allele. The direct H2 tag is primary because it tags H2 directly rather than by subtraction. The joint model is therefore an approximate rather than exact H1-reference parameterisation. Tags were cross-checked against 14 further tagging SNPs (Supplementary Table S1). Plender et al.’s main text transposes the haplogroup labels for rs28542750 and rs769768817 relative to Plender et al. Table S4; we followed the latter (Supplementary methods).

The *MUC5B* H1/H2 discriminator (chr11:1,244,757; H2 r²=1.0) was taken from the Plender main text; being strand-ambiguous, its orientation was resolved by allele frequency (Supplementary methods). All tSNPs passed Hardy-Weinberg testing in controls (minimum p=0.39) and were hard-called with no missingness; their alleles, frequencies and tagging r² are in Supplementary Table S1.

### Covariates and exposure

Five genetic principal components, computed with PLINK v2 from LD-pruned typed-array data (309,453 variants) within the 771 analytical participants themselves, were used in all models. Smoking history and lifetime occupational history were collected at the same structured telephone interview, by an interviewer unaware of case status (Reynolds et al., 2023); ever-smoking and age were covariates in all models. Occupational asbestos exposure was ascertained by Peto job-exposure matrix applied to the occupational history (binary classification); 502 of 771 participants (65.1%) were classified as exposed.

### TOPMed sample

The replication sample comprised 1,961 IPF cases and 3,816 controls of European ancestry from the National Institutes of Health Trans-Omics for Precision Medicine (TOPMed) programme, after restriction to participants with complete covariate data. Cases were diagnosed according to American Thoracic Society/European Respiratory Society criteria at centres in the United States, Europe and Australia and underwent whole-genome sequencing through the TOPMed Whole Genome Sequencing in Familial and Sporadic Idiopathic Pulmonary Fibrosis Study. Controls were unaffected individuals aged 50 years or older selected from the TOPMed Genetic Epidemiology of COPD Study (COPDGene), TOPMed Framingham Heart Study (FHS) and TOPMed Multi-Ethnic Study of Atherosclerosis (MESA). All participants gave written informed consent under IRB-approved protocols for their parent studies. No two participants were related at third degree or closer, and European ancestry was assigned using ancestry-informative principal components (Peljto et al., 2023). *MUC5AC* haplogroups were assigned from the same tagging SNPs, alleles and coding used here; the *MUC5B* H1/H2 discriminator failed genotyping quality control and was not analysed. Models were adjusted for sex, age, smoking status and the first four principal components. Unlike IPFJES, the sample includes women, and all models are therefore adjusted for sex.

### Statistical analysis

Logistic regression (Python 3.12; statsmodels) was performed with IPF case/control status as outcome. Haplogroup counts were entered additively (0/1/2 copies); all models adjusted for age and ever-smoking as established IPF risk factors, PC1–5 (genetic ancestry), and recruitment centre (regression N=771).

The *MUC5AC* models, with H1 as reference in the joint model, were (1a) H3 main effect; (1b) H3 + imputed rs35705950 T count; (1c) joint model with H2 and H3 entered simultaneously; (1d) additionally adjusted for rs35705950. The *MUC5B* models were (2a) H1 main effect and (2b) H1 + rs35705950 T count. rs35705950 was also modelled alone.

Sensitivity analyses, detailed in the Supplementary methods and tabulated there where indicated, comprised rs35705950-stratified models (Table S2), alternative H2 tags (Table S3), genotypic rather than additive fits for H2 and H3, imputed *MUC5AC* exon-31 repeat length (Table S4), an agnostic conditional scan of 109 common well-imputed structural variants across the mucin cluster, imputed from the Bai et al. (2026) long-read assembly panel rather than from the TOPMed panel used for the SNP imputation (Table S5), and conditioning of the H2 tag on the mucin-cluster subset of the conditionally-independent 11p15.5 IPF signals reported by Kousathanas et al. (2025). Nested models were compared by likelihood-ratio test.

The same models were fitted in an independent TOPMed IPF case-control sample (described above); between-sample heterogeneity was assessed by a two-sided z test on the difference in log odds ratios, equivalently Cochran’s Q on one degree of freedom, with standard errors recovered from the reported confidence intervals; where the two samples are pooled, this is a fixed-effect inverse-variance meta-analysis (`code/15_orientation_vs_topmed.py`).

Power for the cluster-wide scan was computed for each of the eight mucin-cluster 11p15.5 signals reported by Kousathanas et al. (their Supplementary Table 5), using the European allele frequency and odds ratio reported there. For each signal, genotypes were simulated under Hardy-Weinberg equilibrium at the reported frequency, the scan model was fitted in the 771 analytical participants to obtain the standard error of the log odds ratio, and power was evaluated analytically at the cluster-wide threshold (α = 0.05/54, the effective number of independent tests; `code/17_cluster_scan_power.py`).

Four primary haplogroup coefficients were tested; p-values are two-sided and uncorrected, and nominal results described as such.

## Results

### Participant characteristics

Table 1 shows the participant characteristics. Cases were slightly older (median 77 vs 76 years, p=0.010); ever-smoking (75.8% vs 69.7%, p=0.066) and occupational asbestos exposure (67.5% vs 62.7%, p=0.182) did not differ significantly. The rs35705950 T allele frequency was 0.33 in cases and 0.12 in controls; 59.7% of cases and 23.1% of controls carried at least one copy, and 6.8% and 1.0% respectively were homozygous (p=9.9×10⁻²⁵ for carriage).

**Table 1.**
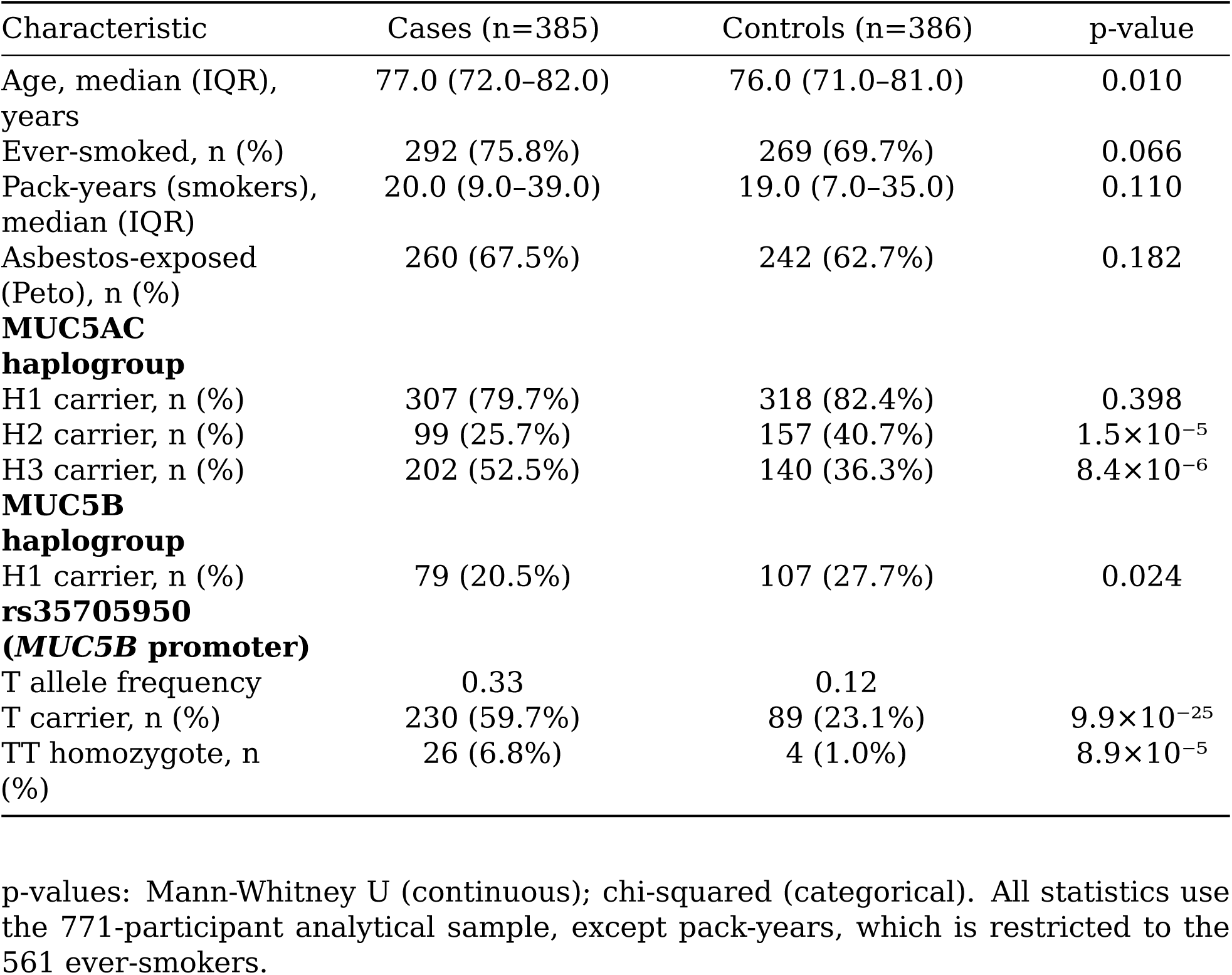
Participant characteristics.

| Characteristic | Cases (n=385) | Controls (n=386) | p-value |
|---|---|---|---|
| Age, median (IQR), years | 77.0 (72.0–82.0) | 76.0 (71.0–81.0) | 0.010 |
| Ever-smoked, n (%) | 292 (75.8%) | 269 (69.7%) | 0.066 |
| Pack-years (smokers), median (IQR) | 20.0 (9.0–39.0) | 19.0 (7.0–35.0) | 0.110 |
| Asbestos-exposed (Peto), n (%) | 260 (67.5%) | 242 (62.7%) | 0.182 |
| **MUC5AC haplogroup** |  |  |  |
| H1 carrier, n (%) | 307 (79.7%) | 318 (82.4%) | 0.398 |
| H2 carrier, n (%) | 99 (25.7%) | 157 (40.7%) | 1.5×10<sup>−5</sup> |
| H3 carrier, n (%) | 202 (52.5%) | 140 (36.3%) | 8.4×10<sup>−6</sup> |
| **MUC5B haplogroup** |  |  |  |
| H1 carrier, n (%) | 79 (20.5%) | 107 (27.7%) | 0.024 |
| **rs35705950 (*MUC5B* promoter)** |  |  |  |
| T allele frequency | 0.33 | 0.12 |  |
| T carrier, n (%) | 230 (59.7%) | 89 (23.1%) | 9.9×10<sup>−25</sup> |
| TT homozygote, n (%) | 26 (6.8%) | 4 (1.0%) | 8.9×10<sup>−5</sup> |

p-values: Mann-Whitney U (continuous); chi-squared (categorical). All statistics use the 771-participant analytical sample, except pack-years, which is restricted to the 561 ever-smokers.

### Haplogroup frequencies

In the IPFJES controls (n=386), which approximate a population reference since cases are enriched for risk haplotypes, haplogroup frequencies were *MUC5AC* H1 ∼57%, H2 ∼23% and H3 ∼20%, and *MUC5B* H2 ∼85%. The *MUC5AC* H3 figure matches the ∼21% reported for Europeans; the *MUC5B* comparator of ∼82% is a panel-wide estimate across all ancestries rather than a European one (Plender et al., 2024). All four haplogroup tags showed no departure from Hardy-Weinberg equilibrium in controls (p=0.45–0.74), as did every variant in the panel (minimum p=0.39; Methods).

### Linkage disequilibrium between *MUC5AC* H3 and rs35705950

Correlations with rs35705950 are reported as the signed r, because the direction of tagging differs between haplogroups; r² is given alongside in Supplementary Table S6. *MUC5AC* H3 count was correlated with rs35705950 T count (r=0.363, p=1.7×10⁻²⁵), sharing ∼13% of its variance; among H3 carriers, 61.4% also carried the T allele. *MUC5B* H1, *MUC5AC* H1 and *MUC5AC* H2 were inversely correlated with T count (r=−0.113, −0.140 and −0.235), so the risk allele co-occurs with the *MUC5B* H2 and *MUC5AC* H3 backgrounds. Carrier status for H3 and for the T allele was nonetheless discordant in 241 of 771 participants (31%). Only H3 has appreciable LD with rs35705950, and its estimate in the IPFJES controls (r²=0.10, slightly lower than the full-sample r² of 0.13 because cases are enriched for risk haplotypes) is consistent with the 1000 Genomes European reference (0.10; Supplementary Table S6), so the LD is representative rather than a feature of this cohort.

### Haplogroup associations with IPF

Full estimates are in Table 2. *MUC5AC* H3 was positively associated with IPF (OR 1.63, 95% CI 1.26–2.09; p=1.8×10⁻⁴), but not independently of rs35705950: adjusting for the promoter variant attenuated H3 to the null (OR 1.10, 95% CI 0.83–1.45; p=0.52), while rs35705950 itself retained a strong effect (OR 3.90, 95% CI 2.83–5.38; p=1.1×10⁻¹⁶). Each additional *MUC5B* H1 copy, in place of H2, was associated with lower odds of IPF (OR 0.71, 95% CI 0.52–0.98; p=0.038), and this likewise attenuated after adjustment (OR 0.82, 95% CI 0.58–1.15; p=0.25).

**Table 2.**
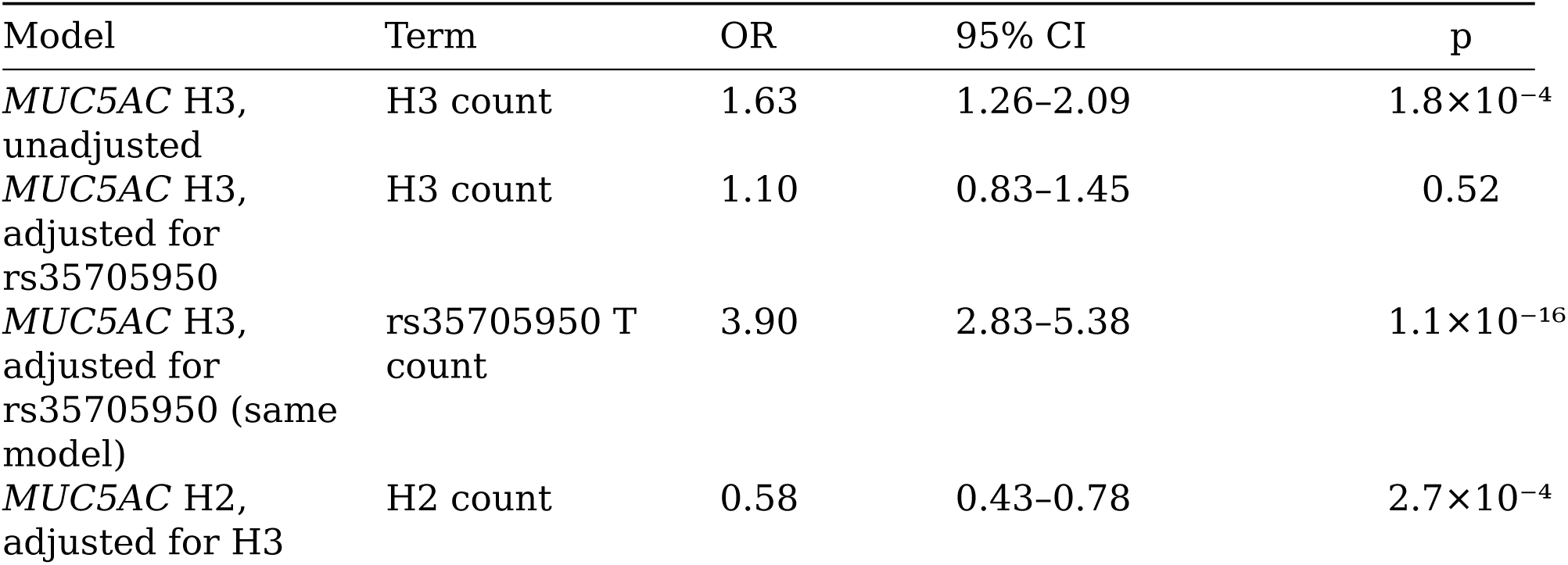
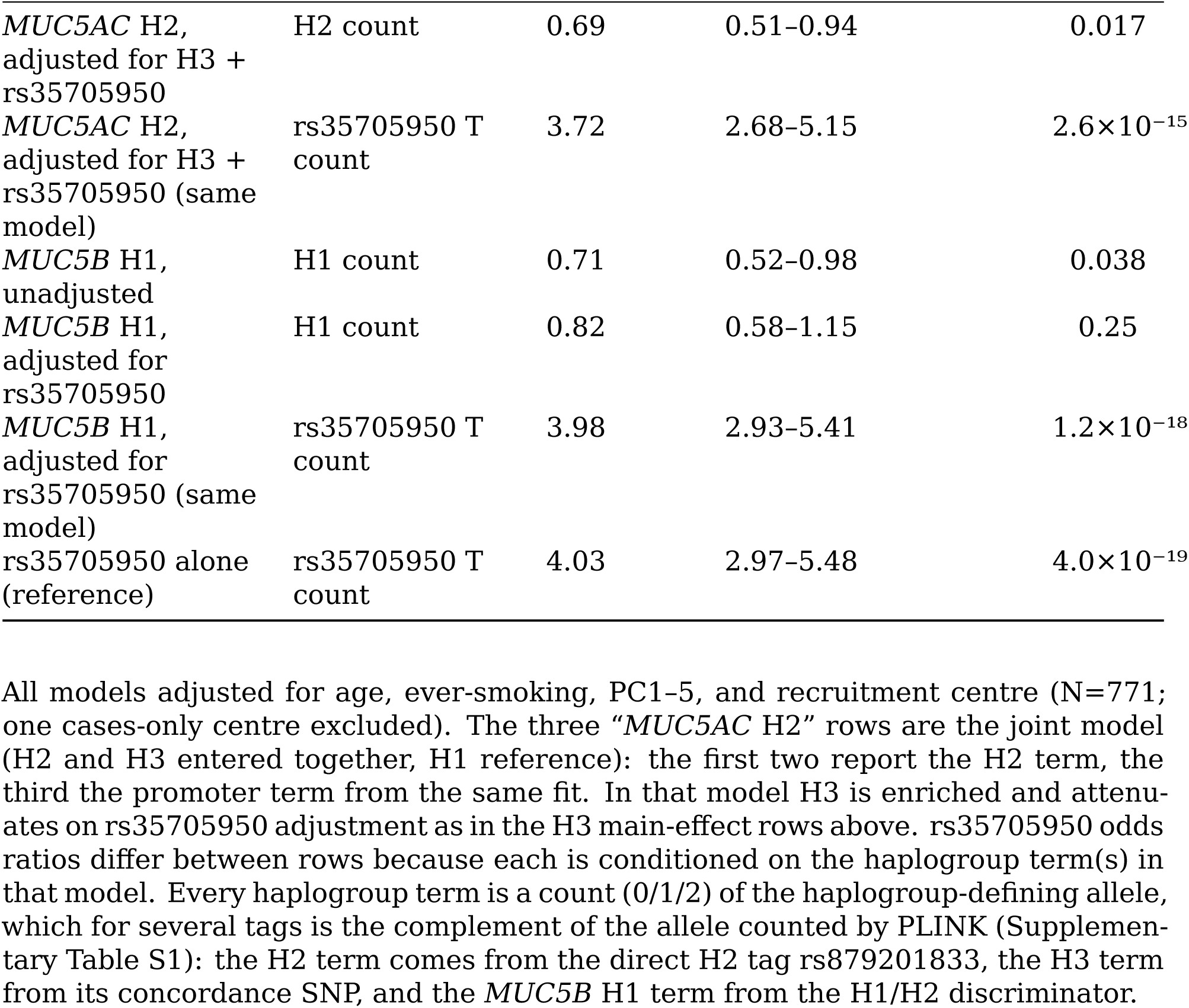
*MUC5AC* and *MUC5B* haplogroup associations with IPF.

| Model | Term | OR | 95% CI | p |
|---|---|---|---|---|
| *MUC5AC* H3, unadjusted | H3 count | 1.63 | 1.26–2.09 | 1.8×10<sup>−4</sup> |
| *MUC5AC* H3, adjusted for rs35705950 | H3 count | 1.10 | 0.83–1.45 | 0.52 |
| *MUC5AC* H3, adjusted for rs35705950 (same model) | rs35705950 T count | 3.90 | 2.83–5.38 | 1.1×10<sup>−16</sup> |
| *MUC5AC* H2, adjusted for H3 | H2 count | 0.58 | 0.43–0.78 | 2.7×10<sup>−4</sup> |
| *MUC5AC* H2, adjusted for H3 + rs35705950 | H2 count | 0.69 | 0.51–0.94 | 0.017 |
| *MUC5AC* H2, adjusted for H3 + rs35705950 (same model) | rs35705950 T count | 3.72 | 2.68–5.15 | 2.6×10<sup>−15</sup> |
| *MUC5B* H1, unadjusted | H1 count | 0.71 | 0.52–0.98 | 0.038 |
| *MUC5B* H1, adjusted for rs35705950 | H1 count | 0.82 | 0.58–1.15 | 0.25 |
| *MUC5B* H1, adjusted for rs35705950 (same model) | rs35705950 T count | 3.98 | 2.93–5.41 | 1.2×10<sup>−18</sup> |
| rs35705950 alone (reference) | rs35705950 T count | 4.03 | 2.97–5.48 | 4.0×10<sup>−19</sup> |

All models adjusted for age, ever-smoking, PC1–5, and recruitment centre (N=771; one cases-only centre excluded). The three “*MUC5AC* H2” rows are the joint model (H2 and H3 entered together, H1 reference): the first two report the H2 term, the third the promoter term from the same fit. In that model H3 is enriched and attenuates on rs35705950 adjustment as in the H3 main-effect rows above. rs35705950 odds ratios differ between rows because each is conditioned on the haplogroup term(s) in that model. Every haplogroup term is a count (0/1/2) of the haplogroup-defining allele, which for several tags is the complement of the allele counted by PLINK (Supplementary Table S1): the H2 term comes from the direct H2 tag rs879201833, the H3 term from its concordance SNP, and the *MUC5B* H1 term from the H1/H2 discriminator.

The three-haplotype joint model (H1 reference) showed the same pattern for H3, but a *MUC5AC* H2 association persisted after additional adjustment for rs35705950, each additional H2 copy in place of H1 being associated with lower odds of IPF (OR 0.69, 95% CI 0.51–0.94; p=0.017, which does not survive correction for the four primary coefficients). This estimate was stable across tag choices, agreeing with the three panel H2 tSNPs and with the diplotype complement, that is, H2 count inferred by subtraction from the other two haplogroups (2 − H1 − H3) rather than from a tag of its own (Supplementary Table S3).

Among participants carrying no copy of the rs35705950 T allele, the promoter variant has no variance and so cannot account for any association. In that stratum H3 was directionally consistent but imprecise (151 cases, 297 controls; OR 1.15, 95% CI 0.79– 1.69; Supplementary Table S2). Genotypic fits for H3 and H2 did not improve on the additive model (Supplementary methods).

### Comparison with an independent TOPMed sample

The same models were fitted in the TOPMed IPF case-control sample (1,961 cases, 3,816 controls; adjusted for sex, age, smoking and PC1–4), with haplogroups assigned from the same tags and coding.

The *MUC5AC* H3 findings replicated: OR 1.62 (95% CI 1.47–1.78) unadjusted in TOPMed against 1.63 (1.26–2.09) here (p=0.96 for heterogeneity). In both samples the association was abolished by adjustment for rs35705950, falling to 0.94 (0.84– 1.05) in TOPMed and 1.10 (0.83–1.45) here (p=0.31 for heterogeneity); neither adjusted interval excludes 1, so this is attenuation to the null rather than a reversal of direction. H2 is estimable only alongside H3, with H1 as reference, so the estimate before adjustment for rs35705950 is the H3-adjusted one: 0.83 (0.74–0.93) in TOPMed against 0.58 (0.43–0.78) here (p=0.028 for heterogeneity). The rs35705950-adjusted H2 residual, however, did not replicate: it was null in TOPMed (OR 0.99, 0.88–1.11), an interval that excludes the 0.69 (0.51–0.94) estimated here, in a sample with five times as many cases. Because IPFJES recruited men only, the TOPMed models were refitted separately by sex (adjusted for age, smoking and PC1–4). The pattern was the same in both: H3 was associated with IPF before adjustment for rs35705950 (men 1.68, 1.48–1.90; women 1.40, 1.19–1.65) and null after it (1.01, 0.88–1.17; 0.85, 0.70–1.02), and the H2 residual was null in both (1.00, 0.87–1.16; 0.97, 0.79–1.19). The failure of the H2 association to replicate is therefore not explained by the male-only composition of IPFJES. The *MUC5B* H1/H2 discriminator failed quality control in TOPMed, so the *MUC5B* association could not be tested there. The samples differ in composition. A higher proportion of TOPMed controls than TOPMed cases had ever smoked (73.9% versus 68.7%), the reverse of the pattern in IPFJES (69.7% of controls versus 75.8% of cases). TOPMed controls came from the COPDGene, Framingham and MESA cohorts. IPFJES recruited men only, and its controls were patients attending outpatient clinics, so neither control group represents the general population.

### Cluster-wide scan of imputed structural variants

11p15.5 contains repeat and structural variation that haplogroup tagging cannot reach, including the *MUC2* VNTR domain. We therefore asked whether any common, well-imputed VNTR or structural-variant allele across the cluster (109 variants) is associated with IPF independently of rs35705950, recognising that this sample can only detect large effects. Twenty-one reached the cluster-wide threshold marginally, but all were attenuated by adjustment for rs35705950 and none reached it after adjustment (Supplementary Table S5). This is a power-limited negative. Taking the effect sizes and allele frequencies Kousathanas et al. report for the eight mucin-cluster 11p15.5 signals, this sample has 74% power to detect their most strongly associated *MUC5B* signal (OR 1.71 at a frequency of 0.20) at the cluster-wide threshold but 1% to 2% power for each of the four *MUC2* and *MUC6* signals, whose odds ratios are 1.15 to 2.10 at allele frequencies of 0.007 to 0.69 (Methods). The absence of those signals here is therefore uninformative.

## Discussion

We tested whether *MUC5AC* or *MUC5B* haplogroup is associated with IPF independently of the *MUC5B* promoter variant rs35705950 in a dedicated IPF case-control study. *MUC5AC* H3 was associated with IPF, but the association is largely explained by its linkage disequilibrium with rs35705950, as was that of *MUC5B* H1. A residual *MUC5AC* H2 association survived adjustment in this cohort but did not replicate in a larger independent sample.

Adjusting rs35705950 for H3 barely changes the rs35705950 estimate (OR 4.03 to 3.90), whereas adjusting H3 for rs35705950 abolishes the H3 estimate (1.63 to 1.10); adding H3 to a model already containing rs35705950 does not improve fit (χ²₁=0.41, p=0.52), whereas the reverse does (χ²₁=77.8, p=1.2×10⁻¹⁸). This pattern is consistent with the H3 association being explained by rs35705950, or by variation more closely tagged by rs35705950 than by the H3 proxy. The LD between them (r=0.36) is the key interpretive constraint: because H3 and rs35705950 are co-inherited, no sample of this size can say whether an H3 association reflects VNTR length, rs35705950-mediated expression, or another co-inherited variant. That LD was also addressed directly rather than only by adjustment: nearly a third of participants are discordant for H3 and T carrier status, and the non-carrier stratum removes the promoter variant’s variance entirely, where H3 remained directionally consistent but imprecise (OR 1.15, 95% CI 0.79–1.69, on 151 cases). The limit there is power, not LD. The tag itself is not the limitation, since it indexes H3 at r²=1.0 in Plender’s panel and tracks independently imputed repeat length at r=0.98 here. The joint-model ordering (H2 depleted, H1 intermediate, H3 enriched for the T allele) is what would be seen if risk tracks rs35705950 frequency by haplogroup rather than repeat length.

*MUC5AC* H2 behaved differently within this cohort: adjustment for rs35705950 did not abolish it (0.58 to 0.69), the estimate was stable across alternative tags and agreed closely with the diplotype complement (r=0.97; Supplementary Table S3), and conditioning on the eight mucin-cluster 11p15.5 signals reported by Kousathanas et al. left the point estimate essentially unchanged, though no longer nominally significant (0.71, p=0.051). However, that residual did not replicate. In the independent TOPMed sample the rs35705950-adjusted estimate was null (OR 0.99, 95% CI 0.88– 1.11), an interval that excludes our estimate, in a sample with five times as many cases, so we do not claim an H2 effect independent of the promoter variant. The samples differ in control ascertainment and in the men-only composition of IPFJES, but the parsimonious reading is that our estimate was inflated by chance in a small sample.

Whether repeat number itself carries risk can be addressed more directly. Imputing *MUC5AC* repeat length into IPFJES from the Bai et al. long-read panel recovered the exon-31 length variants at high fidelity (imputation r²=0.99 for the 751 bp insertion), but imputed length proved equivalent to the clade tag, correlating with H3 count at r=0.98 and carrying the same weak linkage with rs35705950 (r=0.37). It behaved identically to the H3 tag (unadjusted OR 1.60, 95% CI 1.24–2.07; adjusted 1.06, 0.79– 1.41), and a composite of total exon-31 length gave the same result (Supplementary Table S4). The H3 tagging r² of 1.0 is Plender’s panel estimate and need not transfer to a European cohort, but the tag and the independently imputed length agree at r=0.98 in our own data, so little remains for imputation to add. In Bai et al.’s UK Biobank statistics the *MUC5AC* exon-31 repeat is likewise not associated with the pulmonary-fibrosis phecode, an independent and far larger test, though one using a heterogeneous phecode and without conditioning on rs35705950 (Bai et al., 2026).

Cysteine-rich domain copy number and glycosylation density are among the features invisible to a length-based model, as are the predicted protein-truncating haplotypes Plender et al. report at *MUC5B*, whose status is unresolved (Plender et al., 2024). The *MUC5B* H1 inverse association (0.71) is explained as H3’s is: H2, the predominant *MUC5B* haplogroup, carries rs35705950 T, making H1 appear protective by contrast, and the estimate attenuates on adjustment (0.82, p=0.25).

### Strengths and limitations

Strengths include validated tSNPs from long-read sequencing and a cohort recruited specifically for IPF, with MDT-confirmed diagnoses. The principal limitation is that array-based tSNPs index a phylogenetic clade within an LD block, so even a surviving association identifies the clade, not the feature driving it. The second concerns the variants themselves: per-variant imputation quality for the tagging SNPs could not be assessed beyond the r²≥0.5 QC threshold, so imputation error in a tag propagates into haplogroup assignment, and Plender’s tagging r² values are averaged across a 206-haplotype panel assembled from 104 individuals, only 10 of them European, so they carry no guarantee of transferring to a European cohort. The third concerns rs35705950. It lies within the same locus as the haplogroups and in partial linkage disequilibrium with them, so it is not an external covariate: adjusting for it may remove part of the haplogroup signal itself, and the adjusted estimate asks whether haplogroup matters among chromosomes already matched on promoter genotype, which is a narrower question than whether haplogroup matters at all. The fourth is power: where an association attenuates, this sample cannot exclude a modest residual (the adjusted *MUC5AC* H3 interval, 0.83–1.45, remains compatible with a per-allele OR of about 1.45), though pooled with TOPMed it narrows to 0.96 (0.87–1.07). IPFJES recruited men only, so applicability to women is unknown.

### Conclusions

*MUC5AC* H3 and *MUC5B* H1 are associated with IPF risk in directions consistent with their LD with rs35705950, and neither association is separable from it. A *MUC5AC* H2 association did survive adjustment for rs35705950 here, but was not replicated in an independent sample with five times as many cases, where the rs35705950-adjusted estimate was null (OR 0.99, 95% CI 0.88–1.11). We therefore do not claim a haplogroup effect independent of the promoter variant.

## Data Availability

Analysis code, the exact genotype-handling commands and four independent verification layers are available at https://github.com/drcjar/ipfjes-mucin-vntr. Individual-level participant data are held under IPFJES governance arrangements (REC 17/EM/0021) and are available to qualified researchers upon application to the corresponding author and completion of a data access agreement.

## Code and data availability

analysis code, the exact genotype-handling commands (code/REPRODUCE.md), and four independent verification layers are available at https://github.com/drcjar/ipfjes-mucin-vntr. Individual-level participant data are held under IPFJES governance arrangements and are available to qualified researchers upon application to the corresponding author and completion of a data access agreement.

## Supplementary methods: software and reproducibility

Analyses used PLINK v2.00a3.7 for LD-pruning, PCA, dosage export and Hardy-Weinberg testing, and PLINK v1.90b6.26 for single-variant text extractions, with Python 3.12 (numpy, scipy, pandas, statsmodels, pandas-plink) for statistical analysis. Genetic principal components were computed from LD-pruned typed-array data (`--indep-pairwise 500 50 0.2`, then `--pca`); haplogroup-tagging SNP dosages were extracted from imputed chromosome 11 (`--export A`); and tag-SNP Hardy-Weinberg equilibrium and missingness were tested in controls (--hardy). The exact commands, software versions, and the analysis and numerical-verification scripts are provided in the code repository (`code/REPRODUCE.md and code/README.md`).

### Haplogroup label assignment

The haplogroup labels for rs28542750 and rs769768817 in Plender et al.’s main text appear transposed relative to Plender et al. Supplementary Table S4. That table resolves it, giving per-haplogroup r² of 0.111 (H1), 0.055 (H2) and 1.000 (H3) for rs28542750, and 0.899 (H1), 0.012 (H2) and 0.001 (H3) for rs769768817. We therefore assigned rs28542750 to H3 and rs769768817 to H1. Our own control frequencies corroborate this: rs28542750 (0.203) falls within the H3 tag set and matches the∼21% European H3 frequency, and rs769768817 (0.571) within the H1 tag set; this also fixes the orientation of both palindromes. The primary inference is in any case invariant to the labelling; only which clade is described as the long one would change.

### *MUC5B* discriminator orientation

The major allele (0.852 in controls) tags the common H2 haplogroup, and a minorallele frequency of 0.148, far from 0.5, makes the assignment unambiguous despite the palindrome (consistent with Plender’s panel-wide H2 estimate of ∼82%). This was corroborated by the inverse correlation of *MUC5B* H1 with the rs35705950 (chr11:1,219,991 GRCh38) risk allele (r=−0.11; Results), consistent with the T allele being carried predominantly on the common *MUC5B* H2 background.

### Sensitivity analyses

A fifth Plender H2 tag, rs1201597847 (H2 r²=0.859), is present in the imputed data but adds nothing, being in complete LD with the panel tags. rs35705950 was additionally estimated within H3-carrier strata, where it was comparable in H3 non-carriers (OR 3.35, 95% CI 2.11–5.32; N=429) and H3 carriers (4.40, 2.71–7.14; N=342), with no interaction (p=0.42). Because Plender et al. infer heterozygote advantage at H3 from balancing-selection signatures in 1000 Genomes Europeans, *MUC5AC* H3 was also fitted as a two-degree-of-freedom genotypic term, as was *MUC5AC* H2 in the joint model. *MUC5AC* VNTR repeat length was imputed into the 771 from the Bai et al. long-read reference panel using the ImputeSV server (SHAPEIT5 phasing, Minimac4), and both the 751 bp exon-31 length allele and a composite of total exon-31 repeat length were tested against IPF with and without adjustment for rs35705950 (Supplementary Table S4; `code/10_imputed_vntr_length.py`). The cluster-wide scan covered every common (MAF≥0.01) well-imputed (imputation r²≥0.8) VNTR and structural-variant allele across *MUC6*–*MUC5B* (chr11:1.01–1.263 Mb; 109 variants after de-duplicating the VNTR and SV panels), with the cluster-wide threshold set from the effective number of independent tests (Li and Ji, 2005; Supplementary Table S5; `code/11_mucin_cluster_scan.py`). Kousathanas et al. report nine conditionally-independent 11p15.5 signals. We conditioned on the eight lying within the mucin cluster (chr11:1.01–1.263 Mb; by their nearest-transcription-start-site annotation, one nearest *MUC6*, three *MUC2* and four *MUC5B*), retaining rs35705950 in the model; the ninth (rs34787245, chr11:1,289,602) lies in *TOLLIP*, outside the cluster, and was not included (`code/12_h2_conditional_on_locus_signals.py`).

## Supplementary Tables

**Supplementary Table S1.** Haplogroup-tagging SNP panel: alleles, frequencies, and tagging quality. Alleles = reference/alternate on the GRCh38 forward strand, as carried in the TOPMed variant ID. This is not the A1/A2 ordering of our PLINK 1 binary filesets, in which the major allele occupies A2 (PLINK 1.x set major alleles to reference automatically); the two coincide only where the reference allele is also the major allele, 13 of the 19 variants here. Counted = the allele counted in the PLINK dosage. PLINK 2--export A counts the reference allele, which for these filesets is A2, so the counted allele is the major allele throughout and every Freq exceeds 0.5. Freq = its frequency in the IPFJES controls (n=386), which approximate a population reference; MAF = minor-allele frequency, equal to 1 - Freq here. Hap. allele = the allele that defines the tagged haplogroup, which is not always the counted allele: the seven H3 tags, the three *MUC5AC* H2 panel tags, the direct H2 tag and rs35705950 are entered into the models as 2 − dosage, so for those twelve the defining allele is the other allele at the site and its frequency is 1 − Freq. The *MUC5B* discriminator is counted on its H2 allele, so the *MUC5B* H1 term is likewise 2 − dosage. For rs35705950, which is not a haplogroup tag, this column gives the T risk allele. r² (final column) = haplogroup-tagging r² from Plender et al. Supplementary Table S4, except rs35779873, whose r²=0.805 is from Plender et al. Table S6 (it is absent from their Table S4, being below their stated r²>0.85 discovery threshold, but Plender list it among the *MUC5AC* H3 tSNPs in their Methods). The *MUC5B* discriminator r² is from the Plender main text; rs35705950 is the promoter variant, not a haplogroup tag. Strand-ambiguous (palindromic) C/G or A/T tSNPs had their orientation resolved by allele frequency (Methods), unambiguous where the MAF is away from 0.5, notably the *MUC5B* C/G discriminator (MAF 0.148) and the A/T H3 concordance tag rs28542750 (MAF 0.203, matching the ∼21% European H3 frequency). Plender et al. Table S4 gives GRCh38 locations and tagging r² but no alleles, so the location fixes which variant is meant while leaving open which of its two alleles tags the haplogroup; the TOPMed variant ID supplies the reference and alternate alleles at that site, and the tagging allele was resolved from the risk allele in Plender et al. Table S6 where listed and otherwise by the counted-allele frequency in our controls (Methods). The H1 concordance tag (rs769768817; r²=0.899, MAF 0.43) is used only as a cross-check; in the primary analysis H1 is the reference baseline and is not entered as a dosage term. Computed by `code/05_tsnp_table.py`.

| tSNP | Position | Alleles | Counted | Freq | MAF | Hap. allele | Tags | r<sup>2</sup> |
|---|---|---|---|---|---|---|---|---|
| rs28542750 | chr11:1165501 | T/A | T | 0.797 | 0.203 | A | *MUC5AC* H3 conc. | 1.0 |
| rs769768817 | chr11:1195214 | G/C | C | 0.571 | 0.429 | C | *MUC5AC* H1 conc. | 0.899 |
| rs2075842 | chr11:1193830 | G/C | G | 0.574 | 0.426 | G | *MUC5AC* H1 | 0.923 |
| rs1132433 | chr11:1194354 | A/G | A | 0.574 | 0.426 | A | *MUC5AC* H1 | 0.923 |
| rs1132434 | chr11:1195265 | G/C | C | 0.573 | 0.427 | C | *MUC5AC* H1 | 0.923 |
| rs28652890 | chr11:1195858 | T/C | T | 0.573 | 0.427 | T | *MUC5AC* H1 | 0.923 |
| rs879136008 | chr11:1182048 | A/T | T | 0.574 | 0.426 | T | *MUC5AC* H1 | 0.923 |
| rs28519516 | chr11:1202343 | T/C | C | 0.773 | 0.227 | T | *MUC5AC* H2 | 0.862 |
| rs28558973 | chr11:1202859 | A/G | G | 0.773 | 0.227 | A | *MUC5AC* H2 | 0.862 |
| rs28368633 | chr11:1202914 | T/C | C | 0.773 | 0.227 | T | *MUC5AC* H2 | 0.862 |
| rs36154966 | chr11:1178216 | C/G | C | 0.803 | 0.197 | G | *MUC5AC* H3 | 1.0 |
| rs1004828576 | chr11:1178023 | A/G | A | 0.797 | 0.203 | G | *MUC5AC* H3 | 1.0 |
| rs940158763 | chr11:1177688 | A/T | A | 0.797 | 0.203 | T | *MUC5AC* H3 | 1.0 |
| rs36151150 | chr11:1177356 | T/C | T | 0.797 | 0.203 | C | *MUC5AC* H3 | 1.0 |
| rs36132281 | chr11:1177269 | T/C | T | 0.797 | 0.203 | C | *MUC5AC* H3 | 1.0 |
| rs35779873 | chr11:1197482 | C/T | C | 0.799 | 0.201 | T | *MUC5AC* H3 | 0.805 |
| rs35705950 | chr11:1219991 | G/T | G | 0.880 | 0.120 | T | *MUC5B* promoter | n/a |
| rs879201833 | chr11:1182279 | A/G | A | 0.775 | 0.225 | G | *MUC5AC* H2 direct | 0.888 |
| *MUC5B* discriminator | chr11:1244757 | C/G | C | 0.852 | 0.148 | C | *MUC5B* H2 | 1.0 |

**Supplementary Table S2.** *MUC5AC* H3, *MUC5AC* H2 and *MUC5B* H1 associations stratified by rs35705950 carrier status. All models adjusted for age, ever-smoking, PC1–5, and recruitment centre. Noncarrier stratum excludes centre 18 (n=4, cases only); carrier stratum excludes centres 17 (n=2) and 21 (n=4) (cases only), to avoid complete separation. H3 OR in non-carriers (1.15) is consistent with confounding but the analysis is underpowered to exclude an independent effect of modest magnitude. *MUC5AC* H2 rows are from the joint model (H2 tag and H3 entered together, H1 reference). The “Overall” rows are not uniform: for H3 and *MUC5B* H1 the estimate is the unadjusted single-term one, whereas for H2 it is the rs35705950-adjusted joint estimate (0.69; Table 2), shown because the H2 estimate of interest is the one adjusted for rs35705950, which is the estimate that did not replicate in TOPMed (Results). The non-carrier rows control for rs35705950 by design, since the promoter variant has no variance in that stratum; the carrier rows pool T=1 and T=2 without adjusting for T count.

| Haplogroup | Stratum | N | OR | 95% CI | p |
|---|---|---|---|---|---|
| *MUC5AC* H3 | Non-carrier (T=0) | 448 | 1.15 | 0.79–1.69 | 0.46 |
| *MUC5AC* H3 | Carrier (T≥1) | 313 | 1.00 | 0.64–1.54 | 0.98 |
| *MUC5AC* H3 | Overall | 771 | 1.63 | 1.26–2.09 | 1.8×10<sup>−4</sup> |
| *MUC5B* H1 | Non-carrier (T=0) | 448 | 0.72 | 0.46–1.11 | 0.14 |
| *MUC5B* H1 | Carrier (T≥1) | 313 | 1.03 | 0.54–1.95 | 0.93 |
| *MUC5B* H1 | Overall | 771 | 0.71 | 0.52–0.98 | 0.038 |
| *MUC5AC* H2 | Non-carrier (T=0) | 448 | 0.69 | 0.48–1.01 | 0.054 |
| *MUC5AC* H2 | Carrier (T≥1) | 313 | 0.54 | 0.30–0.97 | 0.040 |
| *MUC5AC* H2 | Overall | 771 | 0.69 | 0.51–0.94 | 0.017 |

**Supplementary Table S3.** *MUC5AC* H2 association across tagging measures. Ctrl freq = frequency in the IPFJES controls (n=386) of the H2-defining allele, or of H2 itself for the complement; r (compl.) = correlation with the diplotype complement. The primary H2 measure is the direct tag rs879201833 (r²=0.888). The three Plender panel tags are in complete mutual linkage disequilibrium in our data (tag-to-tag r=1.000) and give identical estimates, so they are shown as a single row; each was nonetheless fitted separately and all four estimates are verified individually by `code/verify_numbers.py`; the diplotype complement (2 − H1 − H3) is shown as a cross-check. All measures agree in direction and magnitude (adjusted OR 0.66–0.73), so the H2 residual is not an artefact of how H2 is assigned; the two tag sets have comparable tagging quality (Plender r² 0.888 and 0.862; the complement is derived by subtraction and has no tagging r² of its own), and the small differences between the three measures should not be over-read. All models are the *MUC5AC* joint model (H2 and H3 entered together, H1 reference), adjusted for age, ever-smoking, PC1–5 and recruitment centre (N=771). Computed by `code/09_h2_direct_and_lrt.py`.

| H2 measure | Plender r<sup>2</sup> | Ctrl freq | r (compl.) | Unadjusted OR (95% CI) | +rs35705950 OR (95% CI) | p |
|---|---|---|---|---|---|---|
| rs879201833 (primary) | 0.888 | 0.225 | 0.972 | 0.58 (0.43–0.78) | 0.69 (0.51–0.94) | 0.017 |
| Plender panel tags (rs28519516, rs28558973, rs28368633) | 0.862 | 0.227 | 0.968 | 0.55 (0.41–0.74) | 0.66 (0.49–0.90) | 0.009 |
| Diplotype complement (2 − H1 − H3) | n/a | 0.225 | 1.000 | 0.63 (0.47–0.83) | 0.73 (0.54–0.98) | 0.038 |

**Supplementary Table S4.**
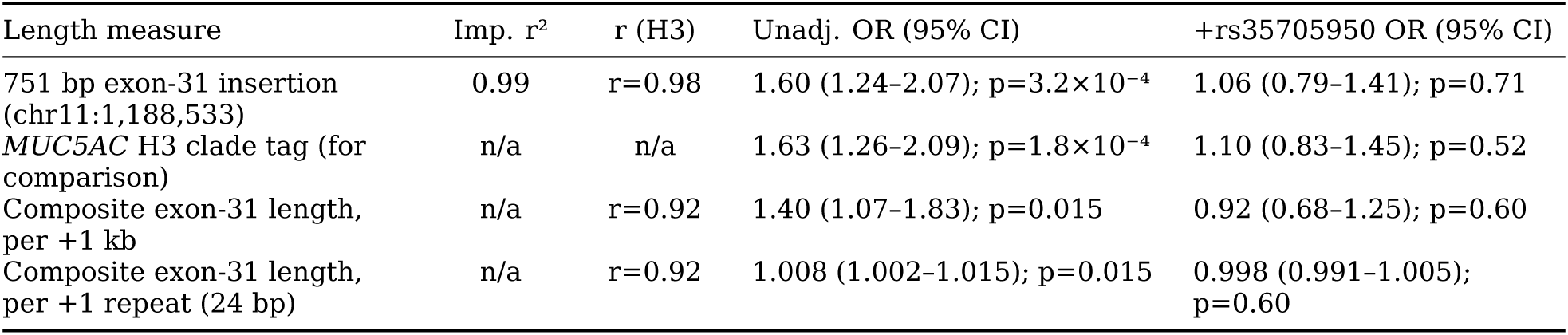
Imputed *MUC5AC* exon-31 repeat length versus IPF. Imp. r² = imputation r²; r (H3) = correlation with imputed H3 count. Repeat lengths were imputed into the 771 analytical participants from the Bai et al. (2026) long-read VNTR reference panel using the ImputeSV server (BCFtools QC → SHAPEIT5 v5.0.1 phasing → Minimac4 v4.1.6); the 751 bp allele is the exon-31 length variant that tracks the H3 clade in long-read assemblies. Imputation r² is the server’s estimate. The composite is total inserted exon-31 sequence per participant relative to GRCh38, summed across imputed alleles (r²≥0.30); results are unchanged if built from the two common well-imputed alleles alone (r=0.96 between definitions). Adding H3 to a model containing the imputed length, or vice versa, does not improve fit (likelihood-ratio p=0.24 and 0.76): the imputed length and the clade tag are the same information. The imputed length therefore behaves as the H3 tag does, associated unadjusted and abolished by rs35705950, consistent with the paper’s argument that an SNP-imputed length inherits the tags’ linkage disequilibrium and cannot isolate the repeat. Models adjust for age, ever-smoking, PC1–5 and recruitment centre. Computed by `code/10_imputed_vntr_length.py`; submission and provenance in `code/REPRODUCE.md` (step 6).

| Length measure | Imp. r<sup>2</sup> | r (H3) | Unadj. OR (95% CI) | +rs35705950 OR (95% CI) |
|---|---|---|---|---|
| 751 bp exon-31 insertion (chr11:1,188,533) | 0.99 | r=0.98 | 1.60 (1.24–2.07); p=3.2×10<sup>−4</sup> | 1.06 (0.79–1.41); p=0.71 |
| *MUC5AC* H3 clade tag (for comparison) | n/a | n/a | 1.63 (1.26–2.09); p=1.8×10<sup>−4</sup> | 1.10 (0.83–1.45); p=0.52 |
| Composite exon-31 length, per +1 kb | n/a | r=0.92 | 1.40 (1.07–1.83); p=0.015 | 0.92 (0.68–1.25); p=0.60 |
| Composite exon-31 length, per +1 repeat (24 bp) | n/a | r=0.92 | 1.008 (1.002–1.015); p=0.015 | 0.998 (0.991–1.005); p=0.60 |

**Supplementary Table S5.** Agnostic conditional scan of imputed structural variants across the 11p15.5 mucin cluster. Every common, well-imputed VNTR and structural-variant allele in the mucin cluster was tested against IPF by logistic regression (adjusted for age, ever-smoking, PC1–5 and recruitment centre), marginally and with rs35705950 T-allele count added; loci present in both the VNTR and SV panels were de-duplicated and kept at their higher imputation r². The strong marginal associations, concentrated in the *MUC2* VNTR domain that no haplogroup analysis reaches, all reflect linkage disequilibrium with rs35705950 (top hits r=0.24–0.53 with the T allele) and are markedly attenuated by adjustment for it (the strongest, a *MUC2* allele, from OR 2.77 to 1.64); none survives the cluster-wide threshold, and the smallest adjusted p is within chance expectation across the correlated tests. Survival of adjustment would in any case not establish a causal repeat effect, since every variant lies in the same linkage-disequilibrium block (Discussion). Within this sample, then, no imputed mucin structural variant reached significance independently of the promoter variant. (The *MUC5AC* H2 haplogroup tags are SNPs, not structural variants, and so are not among the 109 alleles scanned here; the smallest adjusted p in this table, 0.003, is a *MUC2* allele at the low end of the common range scanned, whereas the residual H2 association discussed in the main text is assessed against a different, less stringent multiplicity family, the four primary haplogroup coefficients, not this 54-test cluster-wide threshold.) This is a power-limited negative, not a claim that no such variant exists: in a far larger IPF meta-analysis, GCTA-COJO resolves conditionally-independent signals annotated by nearest gene to *MUC2* (three) and *MUC6* (one), as well as several in *MUC5B* (Kousathanas et al., 2025), which 385 cases are underpowered to detect (*MUC2* and *MUC6* are canonically gastrointestinal mucins, so these labels are positional and need not imply an airway role for those proteins). The locus is therefore genuinely multi-signal; what the haplogroup analysis shows is more specifically that the *MUC5AC* and *MUC5B haplogroups* are not separable from rs35705950. Computed by `code/11_mucin_cluster_scan.py`; provenance in `code/REPRODUCE.md` (step 6).

|  |  |
|---|---|
| Variants tested (*MUC6*–*MUC5B*, chr11:1.01–1.263 Mb) | 109 common (MAF≥0.01), well-imputed (r<sup>2</sup>≥0.8) VNTR + SV alleles from the Bai et al. (2026) long-read assembly panel, via the ImputeSV server (69 VNTR panel, 40 SV panel) |
| Allele size (length change versus reference) | 44 to 2,863 bp, median 291 bp; 106 of 109 at least 50 bp |
| By gene | *MUC2* 73, *MUC5AC* 15, *MUC5B* 8, intergenic/flanking 13, *MUC6* 0 (imputes poorly) |
| Effective independent tests (Li & Ji, 2005) | M_eff = 54; cluster-wide threshold 0.05/M_eff = 9.3×10<sup>−4</sup> |
| Reaching threshold marginally | 21 (mostly *MUC2*; 2 the already-reported *MUC5AC* exon-31) |
| Strongest marginal | *MUC2* +1479 bp @1,096,722: OR 2.77, p=6×10<sup>−12</sup> (r=0.52 with rs35705950 T) |
| Surviving adjustment for rs35705950 | 0; smallest adjusted p=0.003, expected P(min<0.003) over M_eff=54 = 0.15 |

**Supplementary Table S6.** Linkage disequilibrium between the *MUC5AC*/*MUC5B* haplogroups and rs35705950. Each haplogroup is represented by its concordance tSNP (Methods); r is the Pearson correlation of the tag allele-dosage count with the rs35705950 T-allele count, r² its square. The reference column is the r² between rs35705950 and each tSNP in the 1000 Genomes European panel (phase 3; Ensembl LD API), the panel Plender et al. used for their locus-wide LD analysis; Plender et al. do not themselves tabulate haplogroup–rs35705950 LD. Only *MUC5AC* H3 shows appreciable LD with rs35705950, and the IPFJES-controls value (r²=0.10) is consistent with the reference (0.10), indicating the LD is representative rather than a feature of this cohort; the full-sample estimates are modestly larger because cases are enriched for risk haplotypes. H1, H2 and *MUC5B* H1 have little LD with rs35705950: r² is 0.02 or less in the controls and 0.06 or less in the full sample, the largest being *MUC5AC* H2 at 0.06. All three fall below the ∼0.05 floor beneath which the Ensembl API returns no value for the reference panel. Computed by `code/13_ld_haplogroup_rs35705950.py`.

| Haplogroup (concordance tSNP) | IPFJES controls (n=386), r (r²) | IPFJES full sample (n=771), r (r²) | 1000G EUR reference, r² |
|---|---|---|---|
| *MUC5AC* H1 (rs769768817) | −0.13 (0.02) | −0.14 (0.02) | <0.05 |
| *MUC5AC* H2 (rs879201833) | −0.16 (0.02) | −0.24 (0.06) | <0.05 |
| *MUC5AC* H3 (rs28542750) | +0.31 (0.10) | +0.36 (0.13) | 0.10 |
| *MUC5B* H1 (chr11:1,244,757) | −0.10 (0.01) | −0.11 (0.01) | <0.05 |

